# Associations between multiple immune-response-related proteins and neonatal infection: a proximity extension assay based proteomic study in cord plasma of twins

**DOI:** 10.1101/2024.02.14.24302852

**Authors:** Ruoqing Chen, Weiri Tan, Yeqi Zheng, Feng Wu, Hui Liang, Youmei Chen, Xian Liu, Fang Fang, Rui Zhang, Quanfu Zhang, Xu Chen

## Abstract

**Background:** Given their immature immune system, neonates are highly susceptible to infection, a major cause of neonatal death. However, associations between immune-response-related proteins and risk of neonatal infection have yet been systematically investigated.

**Methods:** We conducted a nested case-control study of 149 twins (60 cases and 89 controls, including 34 pairs of discordant twins), within the Shenzhen Baoan Birth and Twin (SZBBTwin) cohort. Using proximity extension assay of Olink Proteomics, 92 immune-response-related proteins were measured in samples of cord plasma. All twins were followed for a diagnosis of infection from birth until 27 days of age. Wilcoxon rank-sum test was used to determine differentially expressed proteins (DEPs), and multivariable logistic regression was used to assess the associations of the levels of proteins with neonatal infection. The receiver operating characteristic curve was plotted to evaluate the predictive performance of DEPs. Enrichment analysis was performed to annotate potential functions and pathways of DEPs.

**Results:** Five DEPs (ITGA11, FCRL6, DDX58, SH2D1A, and EDAR) were identified for neonatal infection. A higher cord plasma level of integrin alpha 11 (ITGA11) was associated with a higher risk of neonatal infection in both the analyses of all twins and discordant twins. The area under the curve achieved 0.835 for the five DEPs. The identified DEPs were mainly involved in immune function and protein binding, and most of them were enriched in the nuclear factor kappa-B pathway.

**Conclusion:** Multiple immune-response-related proteins in cord plasma, particularly ITGA11, are associated with the risk of neonatal infection.

**Key point:** In this nested case-control study, 92 immune-response-related proteins were measured in cord plasma by proximity extension assay. A higher level of ITGA11 was associated with a higher risk of neonatal infection, in the analyses of all twins and discordant twins.

## BACKGROUND

According to the World Health Organization (WHO), neonatal infection causes over 550,000 neonatal deaths annually [1], accounting for approximately 23% of the overall neonatal mortality worldwide. Among the 93,400 neonatal deaths in China during 2015, about 8.5% were due to infections such as pneumonia, sepsis, and meningitis [2]. Many systems in neonates are still under development and maturation; the immature immune system, particularly, makes them highly vulnerable to pathogens, leading to infections [3].

Maternally acquired antibodies, such as IgG transmitted through placenta and sIgA in breast milk, are one of the important resources in the immune system to fight against infections for the neonates [4]. In addition, proteins related to immune response, such as interleukins and cytokines, are associated with resistance to neonatal infection [5]. However, immune-response-related proteins in relation to neonatal infection have not been comprehensively investigated, primarily due to the limitations of traditional techniques to quantify levels of proteins [6]. Insufficient consideration of confounding factors that may impact both protein levels and risk of infection in the neonatal period is another reason [7]. The twin design could control for shared genetic and environmental confounding factors and thus may enhance the strength of causal evidence in omics research [8].

Utilizing the dual recognition and DNA-coupled technique, the proximity extension assay (PEA) from Olink Proteomics platform could simultaneously measure a broad range of proteins with a small volume of sample [9]. It therefore serves a high-throughput proteomic tool without the antibody cross-reactivity that would occur in conventional protein assays [10].

In this nested case-control study, we aimed to investigate the associations of cord plasma levels of 92 immune-response-related proteins with risk of neonatal infection among twins. We also intended to assess the predictive performance of differentially expressed proteins (DEPs), as well as their potential function and pathways for neonatal infection.

## METHODS

### Study design and participants

This is a nested case-control study embedded in the Shenzhen Baoan Birth & Twin (SZBBTwin) cohort, which was initiated in the Baoan Women’s and Children’s Hospital, Shenzhen, China, since May 5^th^, 2020. In the SZBBTwin cohort, women who had their antenatal registration and delivered through cesarean section in Baoan Women’s and Children’s Hospital were invited, together with their neonates. Exclusion criteria were women with a history of infection with hepatitis B, human immunodeficiency virus, toxoplasma, rubella virus, cytomegalovirus, herpes simplex virus, and other infections, as well as women who delivered at a gestational age of less than 28 weeks or on weekends. Informed consent was given by the mother before delivery. The Ethics Committee of Baoan Women’s and Children’s Hospital has approved this study (LLSC-2021-04-01-12-KS) and the SZBBTwin cohort (LLSC2019-02-18).

Until February 6^th^, 2023, 352 pairs of twin neonates (N=704) had been recruited. Biological samples were collected at delivery, including maternal blood, cord blood, amniotic fluid, tissues of umbilical cord, and placenta. Four neonates without cord blood samples were excluded from subsequent measurements and analyses (Figure 1). We followed the twin neonates until 27 days of age, during which we collected all medical records of hospitalizations, outpatient visits, and emergencies from the hospital information system (HIS) of Baoan Women’s and Children’s Hospital.

**Figure 1.**
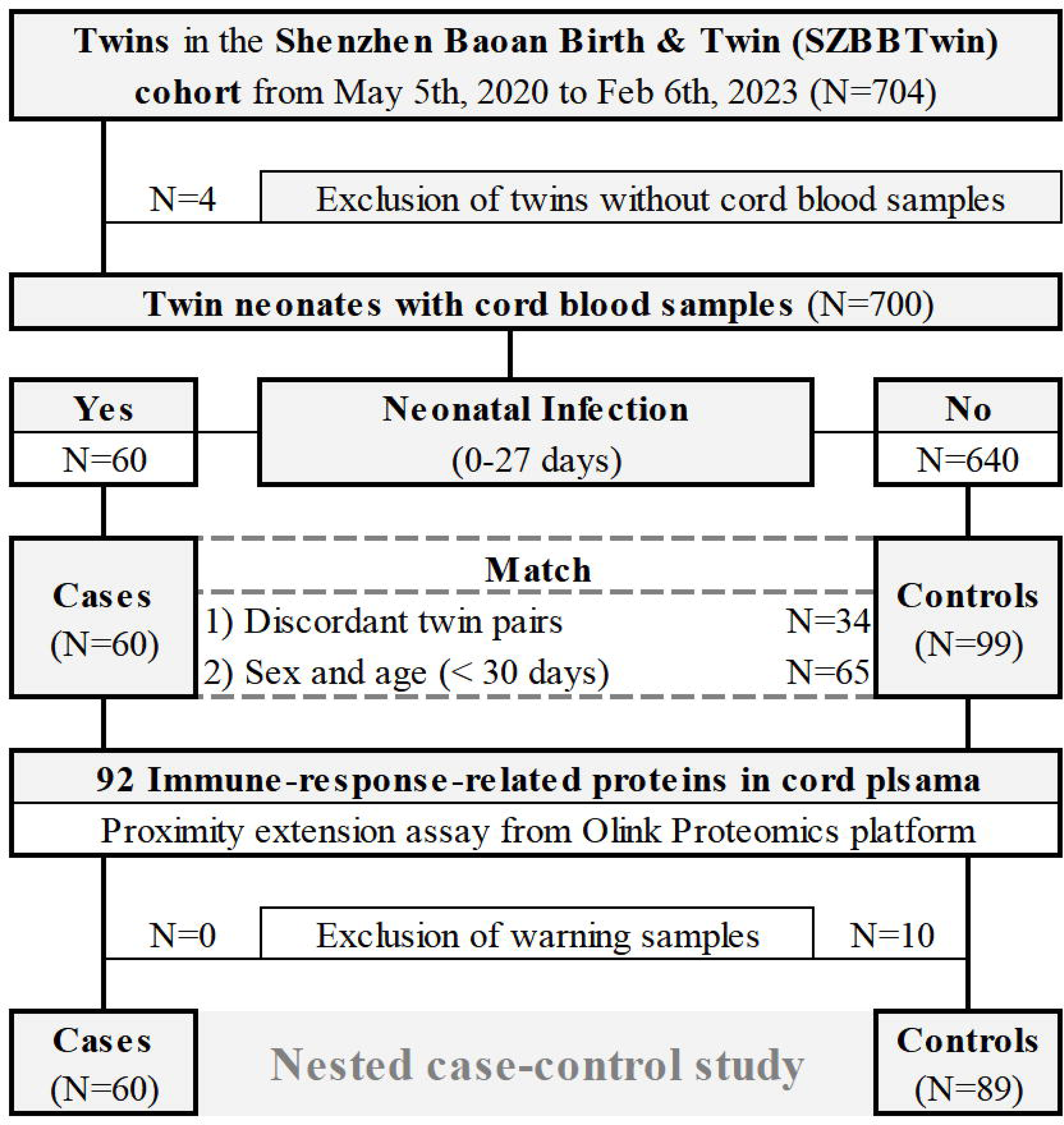
Flow chart of inclusion and exclusion of the study population.

Cases were defined as twin neonates with a clinical diagnosis of infection during 0-27 days after birth. Clinical diagnosis of neonatal infection was determined using diagnostic records in HIS, as well as medical and laboratory tests including blood culture and sputum culture. Discordant twin pairs were defined as pairs in which one twin had neonatal infection, and the other twin had not. A total of 60 cases (including 34 from discordant twin pairs) were identified in the present study, including pneumonia (n=27), acute upper respiratory tract infection (n=11), sepsis (n=5), neonatal conjunctivitis (n=5), coronavirus disease 2019 (COVID-19, n=2), peritonitis (n=2), bronchopneumonia (n=1), acute bronchitis (n=1), meningitis (n=1), perianal abscess (n=1), and other infections (n=4).

Controls were defined as twin neonates who had no infection during 0-27 days after birth. Finally, 99 controls were matched to 60 cases using two strategies (Figure 1): 1) within twin pairs, i.e., discordant twin pairs (n=34); 2) controls and cases other than discordant twin pairs were of the same sex and age (dates of birth less than 30 days apart) (n=65).

### Immune-response-related proteins measurements

Ten milliliters of cord blood were collected with syringes and encapsulated into EDTA vacuum tubes. After centrifugation (4000g) for 15 minutes, the plasma was dispensed into cryotubes and stored at −80℃ until measurements. The entire procedure was completed within six hours.

Using 1µl of each sample of cord plasma, the levels of 92 immune-response-related proteins were measured using PEA (Immune Response Panel, Olink Proteomics AB, Uppsala, Sweden). The assay was performed according to the manufacturer’s instructions and protocol [11], which mainly included four steps: immuno-reaction, extension, pre-amplification, and quantitative real-time polymerase chain reaction. The protein levels measured from PEA were presented as normalized protein expression (NPX) values on a log2-scale, with a higher value corresponding to a higher protein expression. All validation data of the assay (such as detection limits, intra- and inter-assay precision data, etc.) are available on the website of Olink.

Two batches of PEA were performed in our study. Ten plasma samples (including five from cases and five from controls) were used as bridging samples from both batches. Data from two batches were merged and normalized using the R package *OlinkAnalyze* (version 2.0.6). Negative NPX values were retained, and missing NPX values were excluded from the analysis. According to the official recommendation, a warning sample was defined if its principal component (PC) value was 2.5 standard deviations (SD) away from the mean of PC1 and 4 SD away from the mean of PC2. Finally, 10 warning samples were excluded, resulting in 60 cases and 89 controls in our final analysis (Figure 1).

### Confounding factors

Maternal and neonatal characteristics might serve as potential confounding factors for the studied associations. We therefore collected information from the HIS, including maternal age at child’s birth [12], gravidity [13], educational level [14], gestational age [15], vaginal bleeding in early pregnancy [16], pre-pregnancy body mass index (BMI) [17], mode of conception [18], complications included diabetes, hypertension and autoimmune diseases [19–21], as well as sex [22] and birth weight [23] of the neonates.

### Statistical analysis

Continuous variables of prenatal and delivery characteristics were shown as mean and SD, and differences between groups were compared using independent samples t-test or paired samples t-test. Categorical variables were shown as frequencies and proportions, and differences between groups were analyzed using Pearson’s Chi-squared test or Fisher’s exact test. Correlation heatmap and cluster heatmap were used to assess the correlations between different immune-response-related proteins in all twins.

Levels of the immune-response-related proteins between cases and controls were compared using Wilcoxon rank-sum test for all twins, and Wilcoxon signed-rank test for discordant twin pairs. Proteins with statistically significantly different levels (*P*<0.05) among all twins were considered as DEPs. The fold change (FC) values were calculated as the ratio of the means of the NPX values between cases and controls, and the converted FC values and the *P* values from Wilcoxon rank-sum test were used to draw the volcano plot. Logistic regression was used to assess the associations between proteins and risk of neonatal infection in all twins, and conditional logistic regression in discordant twins. Regression models were adjusted for the aforementioned confounding factors, from which the predictive performance was evaluated by the receiver operating characteristic (ROC) curves.

### Enrichment analysis

Metascape was used to perform the enrichment analysis [24], which annotates potential functions, pathways, and interactions for the DEPs, by integrating Gene Ontology (GO) and Kyoto Encyclopedia of Genes and Genomes (KEGG). Terms of enrichment associated with more proteins have smaller *P* values. Term pairs with Kappa similarity exceeding 0.3 are shown as network plots, enabling the visualization of relationships between terms of enrichment.

SAS version 9.4 (SAS Institute, Cary, NC, USA) was used for data management, and R version 4.2.2 (R Foundation for Statistical Computing, Vienna, Austria) was used for statistical analysis and plotting.

## RESULTS

Among all twins in the nested case-control study (n=149), the baseline characteristics showed no difference between cases and controls, except that maternal pre-pregnancy BMI was higher in cases than in controls. Within 34 pairs of discordant twins, cases had higher birth weights than controls (Table 1). The baseline characteristics were not different between all twins and discordant twin pairs (Supplementary Table 1).

**Table 1:**
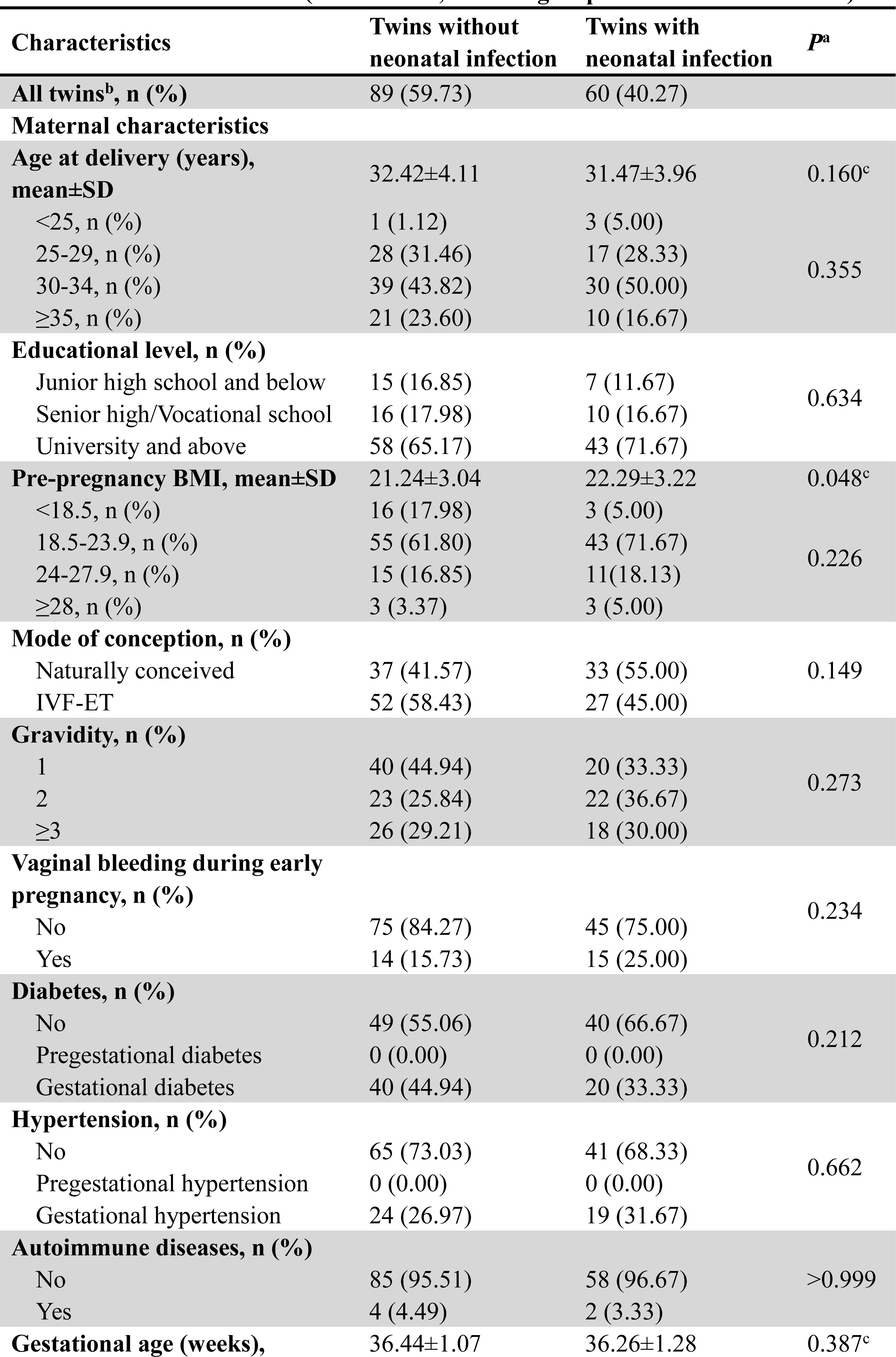
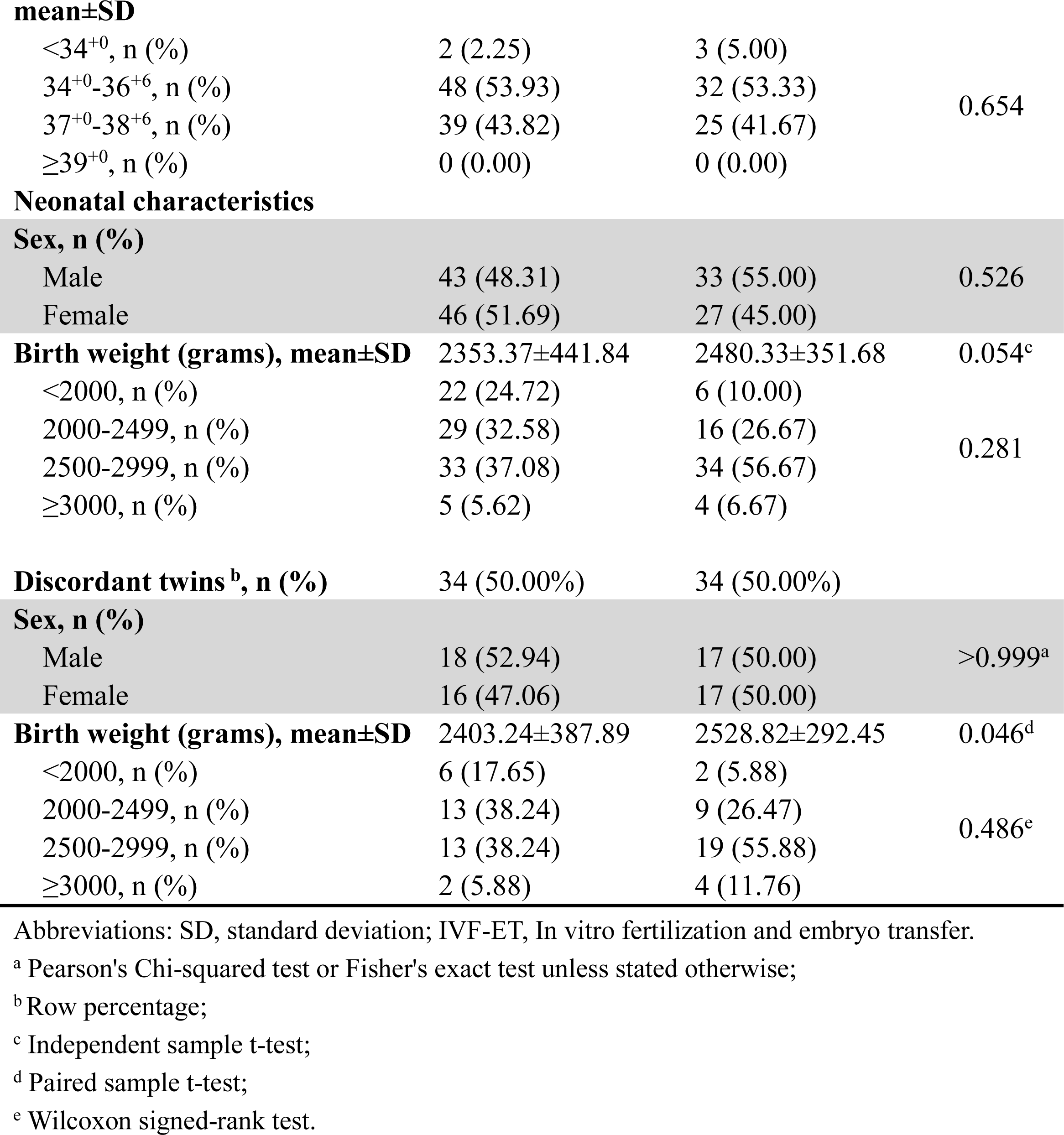
Maternal and neonatal characteristics comparing between twins without and with neonatal infection (in 149 twins, including 34 pairs of discordant twins)

In all twins, cases had higher levels of integrin alpha-11 (ITGA11) and Fc receptor-like 6 (FCRL6), and lower levels of DEAD (Asp-Glu-Ala-Asp) box polypeptide 58 (DDX58), SH2 domain-containing 1A (SH2D1A) and ectodysplasin-A receptor (EDAR), than controls (Figure 2, Supplementary Figure 1). As shown in Supplementary Figure 2, SH2D1A and DDX58 were close to each other on the cluster branches. The correlation heatmap showed a medium to high correlation between DDX58, SH2D1A and EDARs, but a low correlation between ITGA11 and FCRL6 (Supplementary Figure 3). In the discordant twins, cases had lower levels of N-acetylgalactosaminyltransferase 3 (GALNT3), lymphocyte-activation gene 3 (LAG3), and parathyroid hormone receptor (PTH1R) than their co-twins without infection (Supplementary Table 2).

**Figure 2.**
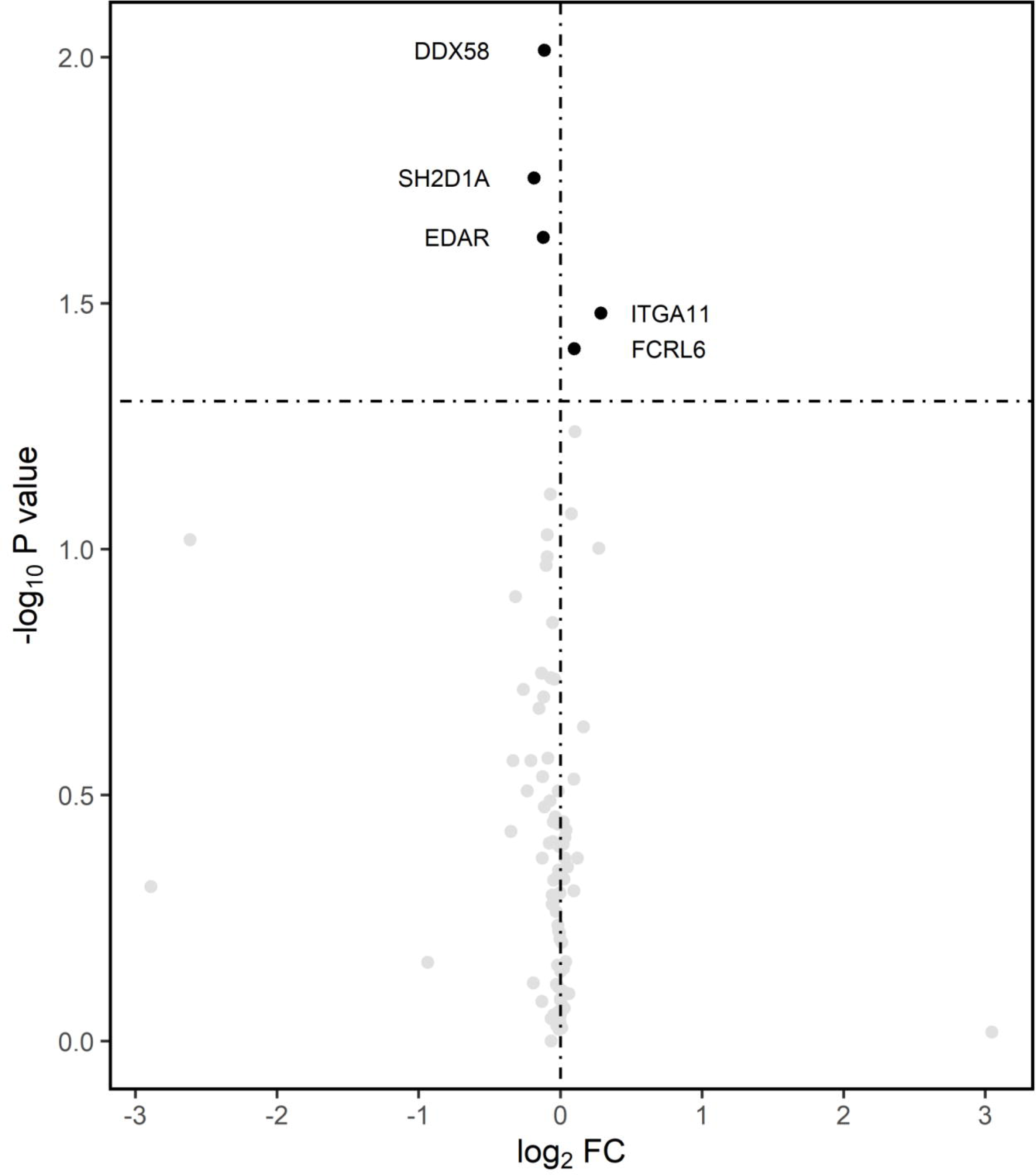
Differentially expressed proteins between neonates with and without infection. FC: fold change values, calculated as the ratio of the means of the normalized protein expression values (case/control). Volcanic plot was used to present the results, in which between-group comparison of levels of proteins showing a P<0.05 (-log_10_P>1.30) from Wilcoxon rank-sum test were defined as differentially expressed proteins.

After multivariable adjustment (Figure 3A, Supplementary Table 3), higher levels of ITGA11, FCRL6, mannose associated serine protease 1 (MASP1), and contactin associated protein-like protein 2 (CNTNAP2) were associated with a higher risk of neonatal infection, whereas inverse associations were observed for SH2D1A, zinc finger and BTB domain-containing 16 (ZBTB16), and sprouty homolog 2 (SPRY2). In the analysis of discordant twin pairs, only higher levels of ITGA11 and proprotein convertase subtilisin/kexin type 4 and serine/arginine-rich splicing factor 1-interacting protein (PSIP1) were associated with a higher risk of neonatal infection (Figure 3B).

**Figure 3.**
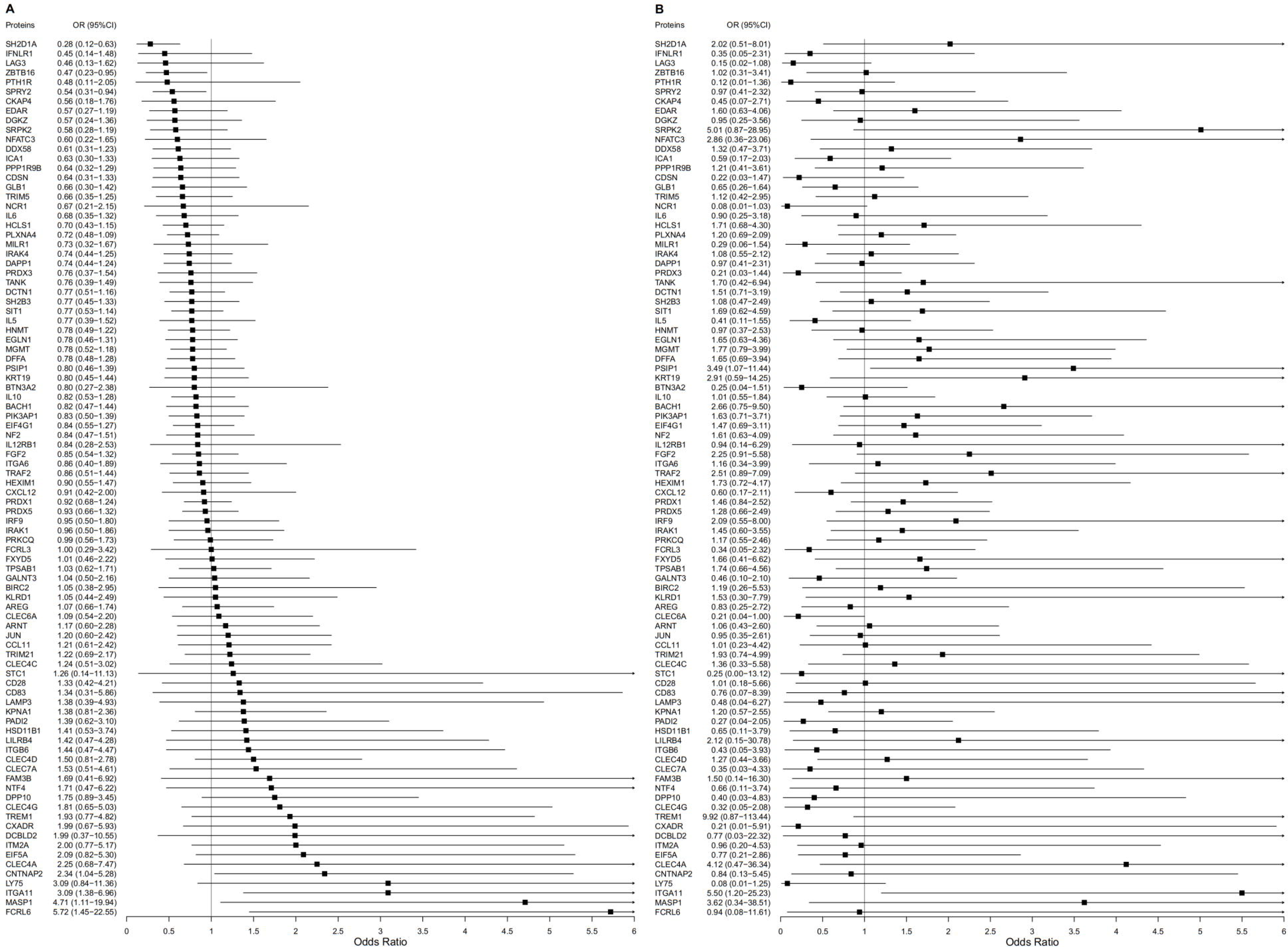
Associations between multiple immune-response-related proteins and neonatal infection. A. In all twins (n=149); B. In discordant twin pairs (n=68, 34 pairs). Forest plot was used to present odds ratio (OR) and 95% confidence interval (CI) from multivariable logistic regression (A) and multivariable conditional logistic regression (B).

The five DEPs (ITGA11, FCRL6, DDX58, SH2D1A, and EDAR) were included one at a time in logistic regression to plot a ROC curve. After the adjustment for confounding factors, the cord plasma levels of the DEPs could predict the probability of neonatal infection with medium accuracy (Figure 4), with the area under curve (AUC) between 0.782 and 0.805 (AUC_ITGA11_ = 0.796, AUC_FCRL6_ = 0.787, AUC_DDX58_ = 0.779, AUC_SH2D1A_= 0.805, AUC_EDAR_ = 0.782, all *P* < 0.001). Furthermore, when the five DEPS were included simultaneously, the regression model showed a specificity of 80.9% and a sensitivity of 75.0% (AUC = 0.835, *P* < 0.001).

**Figure 4.**
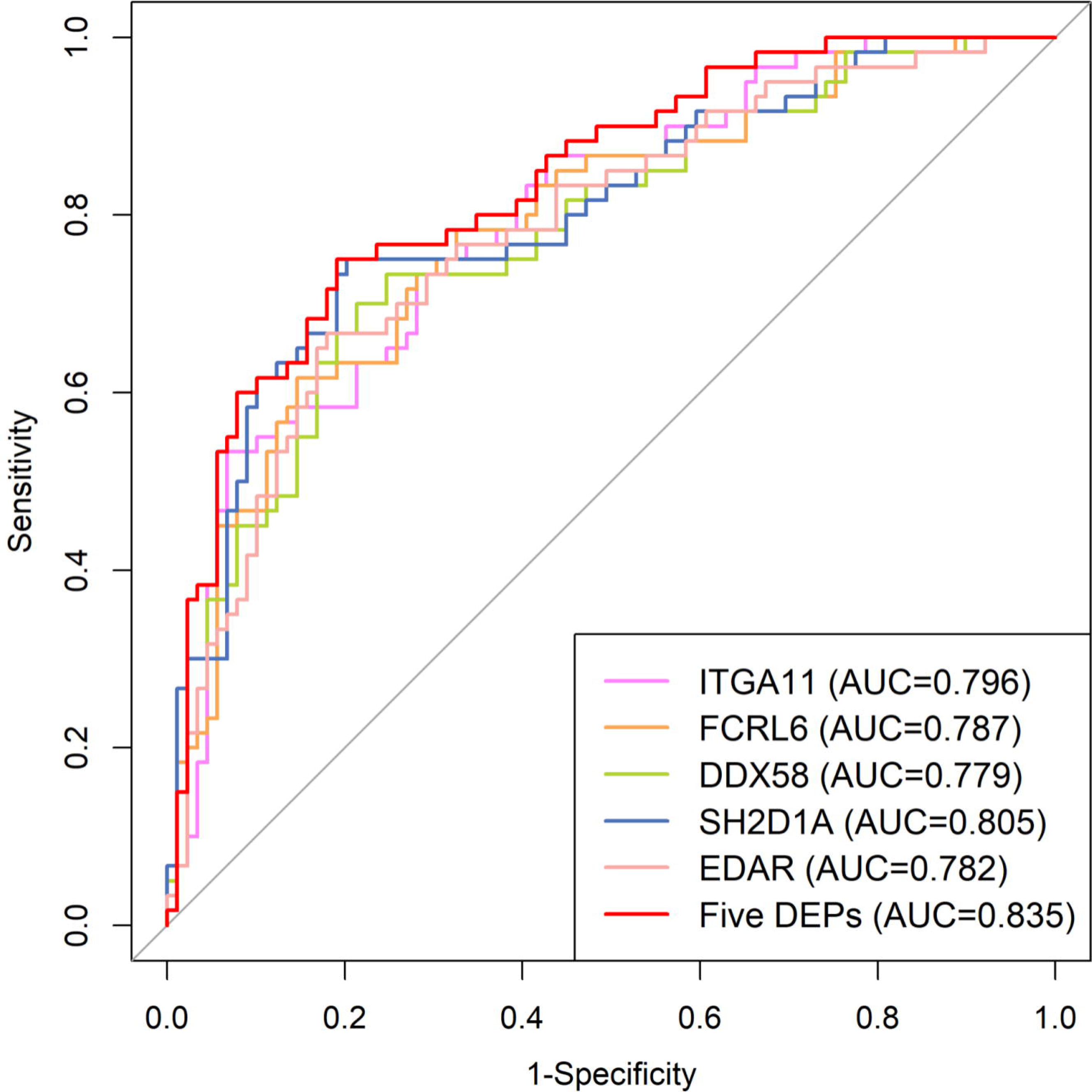
Prediction of differentially expressed proteins for neonatal infection. AUC: Area under the curve.

As shown in the GO enrichment analysis, the five DEPs were mainly enriched in the regulation of leukocyte mediated immunity, integrin alpha11-beta1 complex, regulation of type III interferon production, tube lumen cavitation, and major histocompatibility complex (MHC) class II protein binding (Figure 5A). The network plot (Figure 5B) showed that leukocyte mediated immunity was associated with the regulation of type III interferon production, and integrin alpha11-beta1 complex was associated with MHC class II protein binding (Kappa similarity > 0.3). The KEGG enrichment analysis indicated that the five DEPs were mainly associated with the nuclear factor kappa-B (NF-κB) signaling pathway (Figure 5C).

**Figure 5.**
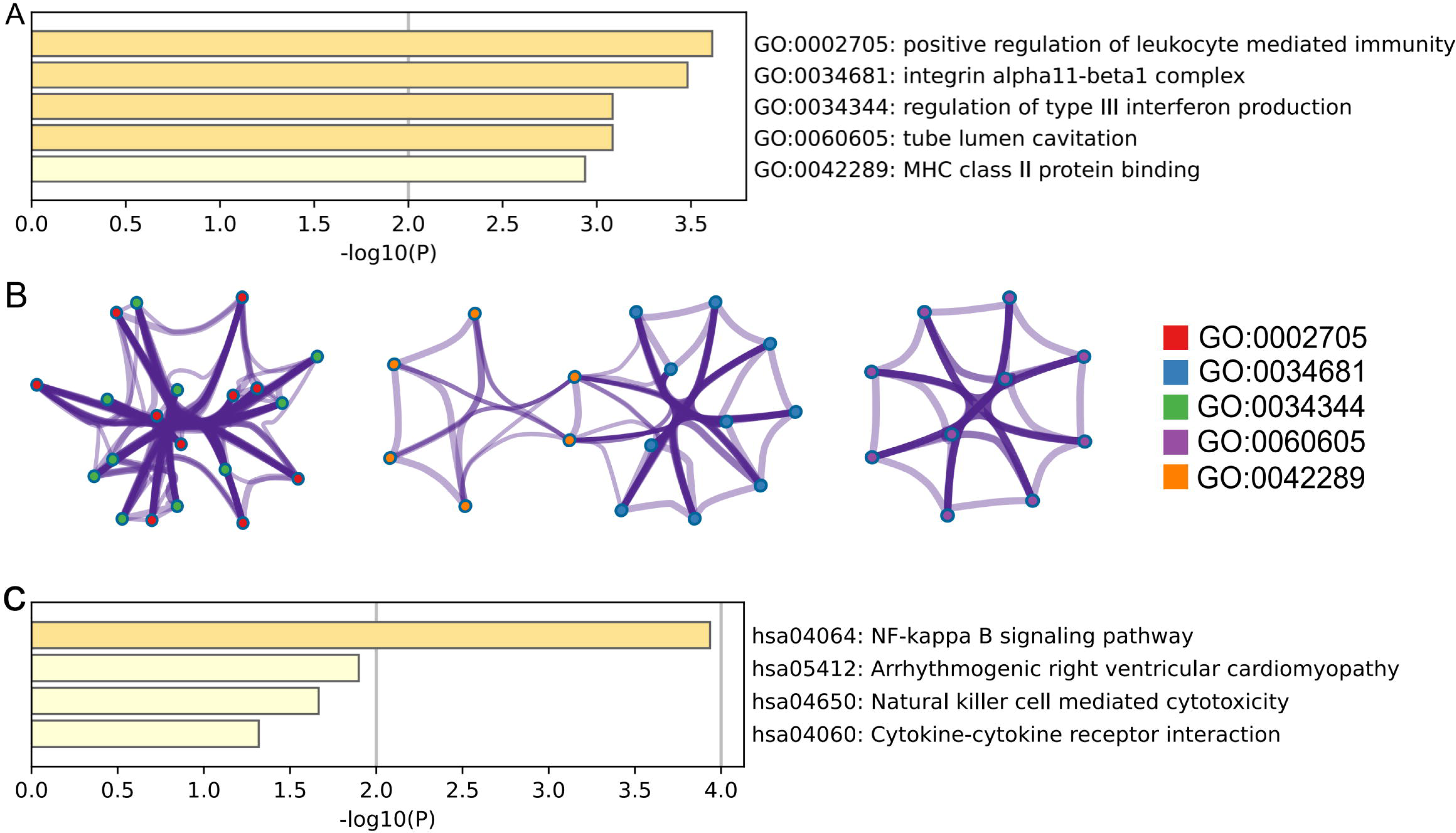
Enrichment analyses for the differentially expressed proteins. A. Bar plot of the Gene Ontology (GO) enrichment; B. Network plot of the GO enrichment; C. Bar plot of the Kyoto Encyclopedia of Genes and Genomes (KEGG) enrichment.

## DISCUSSION

In this nested case-control study of twins, we found that five immune-response-related proteins in cord plasma, particularly ITGA11, were associated with neonatal infection. These five proteins are mainly involved in immune regulation and protein binding, and most of them are enriched in the nuclear factor kappa-B pathway.

Although several immune-response-related proteins have been reported in relation to infection in cell experiments or animal studies [25,26], studies in human neonates are scarce, let alone twins. In our study, cord plasma levels of DDX58, EDAR and SH2D1A were less expressed in cases than controls in all twins. DDX58, an innate immune receptor, has been connected to virus recognition [27] and the initiation of innate immunity and inflammatory responses [28]. As a member of the tumor necrosis factor receptor super-family, EDAR may resist infection and promote inflammation responses by mediating the activation of the NF-κB pathway [29,30]. Previous studies showed that deficiency in SH2D1A was associated with immunodeficiency and might lead to hypogammaglobulinemia and B-cell lymphoma [31,32], corroborating our finding that a lower level of SH2D1A was associated with a higher risk of neonatal infection in all twins. We also observed that a higher level of FCRL6 was associated with a higher risk of neonatal infection. As a receptor for MHC Ⅱ, FCRL6 could regulate humoral responses and may act as a biomarker for chronic immune-related diseases [33]. FCRL6 might also be associated with the reactivation of cytomegalovirus [34].

The five DEPs identified for neonatal infection are enriched in the NF-κB pathway, which may also be one of the main pathways associated with neonatal infection. DDX58 is stimulated during the activation of the non-canonical NF-κB pathway related to RNA viruses infection [35]. SH2D1A can activate NF-κB, which may be important for the host to properly defend against Epstein-Barr virus invasion [36]. The NF-κB pathway participates in the development of various systems in the embryo, and abnormalities in the function of NF-κB pathway can disrupt normal developmental processes or host immune defenses [37].

In the present study, a higher level of ITGA11 was associated with a higher risk of neonatal infection in both the analyses of all twins and discordant twin pairs. ITGA11 is a member of the integrin receptor family, which are dimers composed of α and β subunits and involve in cell-cell or cell-extracellular matrix adhesion. Some studies have reported an association of ITGA11 with infections. In an animal study, ITGA11 was associated with co-infection of avian leukosis virus subgroup J and reticuloendotheliosis virus in chicken embryos, and the study also suggests that ITGA11 was enriched in the pathways of virus-vector interaction, bio-adhesion, and immune responses at the protein level [38]. Similarly, our results of enrichment analysis also indicated that ITGA11 was relevant to protein binding. Binding to cells via integrins might be the infection mechanism employed by many pathogens. For instance, the Arg-Gly-Asp (RGD) tripeptide motif on the envelope of adenoviruses and herpesviruses, which plays a pivotal role in viral infection, acts as a recognition motif for integrins, including ITGA11 [39].

Our study identified several potential protein markers for understanding the immunity to infections among neonates. Pair design of discordant twins was used to control for the effect of shared environmental and genetic confounders, improving the validity of the results, which might also explain the inconsistency in the results of multivariable logistic regressions for all twins and discordant twins [40]. Nested case-control study with a clear chronological order also provides good control of selection and recall bias. In addition, a powerful proteomics technique, i.e., PEA with high-throughput, high-precision, and low-interference, allowed a comprehensive protein analysis.

Our study also has several limitations. We neither performed analyses for different types of neonatal infection (e.g., bacterial, viral, etc.), nor did we subdivide discordant twins into monozygotes and dizygotes, owing to the small sample. We had not performed functional validation due to financial constraints and the scarcity of cord blood of twin neonates. In addition, we analyzed a fixed panel of 92 immune-response-related proteins, and there might be other potentially relevant proteins that had not been included.

In conclusion, five immune-response-related proteins were identified as DEPs for neonatal infection, which might be enriched in the NF-κB pathway. A higher level of ITGA11 was associated with a higher risk of neonatal infection in all twins and discordant twin pairs. The potential role of ITGA11 in the clinical practice of neonatal infection, as well as the molecular mechanisms require further investigation.

## Data Availability

All data produced in the present study are available upon reasonable request to the authors

## ACKNOWLEDGEMENTS

The authors would like to thank Dr. Likuan Xiong, Dr. Bin Liu, Dr. Yuanfang Zhu, and Dr. Xu Chen (Former president) in Shenzhen Baoan Women’s and Children’s Hospital for the construction of the SZBBTwin cohort.

## Author contributions

Ruoqing Chen and Xu Chen conceptualized the study. Weiri Tan, Yeqi Zheng, and Feng Wu retrieved and organized data. Hui Liang performed the assays. Xian Liu, Youmei Chen, Rui Zhang, and Quanfu Zhang recruited the participants and collected the samples. Weiri Tan performed the data collection, processing, and analyses, and wrote the initial draft of the manuscript. Ruoqing Chen, Fang Fang, and Xu Chen were involved in revising the study and manuscript. All authors have read and agreed to the final version of the manuscript.

## Supplementary Data

Supplementary materials are available online.

## References

1. World Health Organization. Newborn infections. Available at: https://www.who.int/teams/maternal-newborn-child-adolescent-health-and-ageing/newborn-health/newborn-infections. Accessed 5 November 2022.

2. He C, Liu L, Chu Y, et al. National and subnational all-cause and cause-specific child mortality in China, 1996–2015: a systematic analysis with implications for the sustainable development goals. Lancet Glob Health 2017; 5:e186–e197.

3. Cinicola B, Conti MG, Terrin G, et al. The protective role of maternal immunization in early life. Front Pediatr 2021; 9:638871.

4. Victora CG, Bahl R, Barros AJD, et al. Breastfeeding in the 21st century: epidemiology, mechanisms, and lifelong effect. Lancet 2016; 387:475–490.

5. Green EA, Garrick SP, Peterson B, et al. The role of the interleukin-1 family in complications of prematurity. Int J Mol Sci 2023; 24:2795.

6. Pla-Roca M, Leulmi RF, Tourekhanova S, et al. Antibody colocalization microarray: a scalable technology for multiplex protein analysis in complex samples. Mol Cell Proteomics 2012; 11:M111.011460.

7. Sly PD, Trottier B, Ikeda-Araki A, Vilcins D. Environmental impacts on infectious disease: a literature view of epidemiological evidence. Ann Glob Health 88:91.

8. Hasegawa M, Taniguchi J, Ueda H, Watanabe M. Twin study: genetic and epigenetic factors affecting circulating adiponectin levels. J Clin Endocrinol Metab 2023; 108:144–154.

9. Coral DE, Franks PW. Proteogenomic mapping sets stage for precision medicine. Nat Metab 2023; 5:366–367.

10. Petrera A, von Toerne C, Behler J, et al. Multiplatform approach for plasma proteomics: complementarity of Olink proximity extension assay technology to mass spectrometry-based protein profiling. J Proteome Res 2021; 20:751–762.

11. Assarsson E, Lundberg M, Holmquist G, et al. Homogenous 96-plex PEA immunoassay exhibiting high sensitivity, specificity, and excellent scalability. PLoS One 2014; 9:e95192.

12. Attali E, Yogev Y. The impact of advanced maternal age on pregnancy outcome. Best Pract Res Clin Obstet Gynaecol 2021; 70:2–9.

13. Lao TT, Sahota DS. Pregnancy and maternal chronic hepatitis B infection—Evidence of reproductive advantage? Am J Reprod Immunol 2017; 77:e12667.

14. Noghanibehambari H, Salari M, Tavassoli N. Maternal human capital and infants’ health outcomes: Evidence from minimum dropout age policies in the US. SSM Popul Health 2022; 19:101163.

15. Miller JE, Hammond GC, Strunk T, et al. Association of gestational age and growth measures at birth with infection-related admissions to hospital throughout childhood: a population-based, data-linkage study from Western Australia. Lancet Infect Dis 2016; 16:952–961.

16. McPherson JA, Odibo AO, Shanks AL, Roehl KA, Macones GA, Cahill AG. Adverse outcomes in twin pregnancies complicated by early vaginal bleeding. Am J Obstet Gynecol 2013; 208:56.e1–56.e5.

17. Rajappan A, Pearce A, Inskip HM, et al. Maternal body mass index: relation with infant respiratory symptoms and infections. Pediatr Pulmonol 2017; 52:1291–1299.

18. Fauser BCJM, Devroey P, Diedrich K, et al. Health outcomes of children born after IVF/ICSI: a review of current expert opinion and literature. Reprod Biomed Online 2014; 28:162–182.

19. De Carolis S, Moresi S, Rizzo F, et al. Autoimmunity in obstetrics and autoimmune diseases in pregnancy. Best Pract Res Clin Obstet Gynaecol 2019; 60:66–76.

20. Bello NA, Zhou H, Cheetham TC, et al. Prevalence of hypertension among pregnant women when using the 2017 American College of Cardiology/American Heart Association blood pressure guidelines and association with maternal and fetal outcomes. JAMA Netw Open 2021; 4:e213808.

21. Ye W, Luo C, Huang J, Li C, Liu Z, Liu F. Gestational diabetes mellitus and adverse pregnancy outcomes: systematic review and meta-analysis. BMJ 2022; 377:e067946.

22. Baines KJ, West RC. Sex differences in innate and adaptive immunity impact fetal, placental, and maternal health. Biol Reprod 2023; 109:256–270.

23. Sahu P, Raj Stanly EA, Simon Lewis LE, Prabhu K, Rao M, Kunhikatta V. Prediction modelling in the early detection of neonatal sepsis. World J Pediatr 2022; 18:160–175.

24. Zhou Y, Zhou B, Pache L, et al. Metascape provides a biologist-oriented resource for the analysis of systems-level datasets. Nat Commun 2019; 10:1523.

25. Liu G, Lu Y, Thulasi Raman SN, et al. Nuclear-resident RIG-I senses viral replication inducing antiviral immunity. Nat Commun 2018; 9:3199.

26. Leomil Coelho LF, Mota BEF, Sales PCM, et al. Integrin alpha 11 is a novel type I interferon stimulated gene. Cytokine 2006; 33:352–361.

27. Sharma A, Kontodimas K, Bosmann M. The MAVS immune recognition pathway in viral infection and sepsis. Antioxid Redox Signal 2021; 35:1376–1392.

28. Loo Y-M, Gale M. Immune signaling by RIG-I-like receptors. Immunity 2011; 34:680–692.

29. Kumar A, Eby MT, Sinha S, Jasmin A, Chaudhary PM. The ectodermal dysplasia receptor activates the nuclear factor-κB, JNK, and cell death pathways and binds to ectodysplasin A. J Biol Chem 2001; 276:2668–2677.

30. Sisto M, Barca A, Lofrumento DD, Lisi S. Downstream activation of NF-κB in the EDA-A1/EDAR signalling in Sjögren’s syndrome and its regulation by the ubiquitin-editing enzyme A20. Clin Exp Immunol 2016; 184:183–196.

31. Tangye SG. XLP: Clinical features and molecular etiology due to mutations in SH2D1A encoding SAP. J Clin Immunol 2014; 34:772–779.

32. Baxter SK, Walsh T, Casadei S, et al. Molecular diagnosis of childhood immune dysregulation, polyendocrinopathy and enteropathy, and implications for clinical management. J Allergy Clin Immunol 2022; 149:327–339.

33. Schreeder DM, Cannon JP, Wu J, Li R, Shakhmatov MA, Davis RS. FCRL6 is an MHC class II receptor. J Immunol 2010; 185:23–27.

34. Adam Alexandersson, Mikko S Venäläinen, Nelli Heikkilä, et al. Proteomics screening post pediatric allogeneic hematopoietic stem cell transplantation reveals an association between increased expression of inhibitory receptor FCRL6 on γδ T cells and CMV reactivation. medRxiv 2023;: 2023.11.02.23297952.

35. Sun S-C. The non-canonical NF-κB pathway in immunity and inflammation. Nat Rev Immunol 2017; 17:545–558.

36. Sylla BS, Murphy K, Cahir-McFarland E, Lane WS, Mosialos G, Kieff E. The X-linked lymphoproliferative syndrome gene product SH2D1A associates with p62dok (Dok1) and activates NF-κB. Proc Natl Acad Sci U S A 2000; 97:7470–7475.

37. Liu P, Li Y, Wang W, et al. Role and mechanisms of the NF-ĸB signaling pathway in various developmental processes. Biomed Pharmacother 2022; 153:113513.

38. Cui X, Zhang X, Xue J, Yao Y, Zhou D, Cheng Z. TMT-based proteomic analysis reveals integrins involved in the synergistic infection of reticuloendotheliosis virus and avian leukosis virus subgroup J. BMC Vet Res 2022; 18:131.

39. Hussein HAM, Walker LR, Abdel-Raouf UM, Desouky SA, Montasser AKM, Akula SM. Beyond RGD: virus interactions with integrins. Arch Virol 2015; 160:2669–2681.

40. McGue M, Osler M, Christensen K. Causal inference and observational research: the utility of twins. Perspect Psychol Sci 2010; 5:546–556.

