## Supplement for "Associations between multiple immune-response-related proteins and neonatal infection: a proximity extension assay based proteomic study in cord plasma of twins"

Supplementary Table 1. Maternal and neonatal characteristics comparing between all twins and discordant twins

Supplementary Table 2. Normalized protein expression values of immune-response-related proteins between neonates without and with infection

Supplementary Table 3. Association between immune-response-related proteins levels and neonatal infection in twins

Supplementary Figure 1. Box plot for proteins levels between infant with or without neonatal infection

Supplementary Figure 2. Cluster heatmap of the levels of multiple immune-response-related proteins

Supplementary Figure 3. Correlation heatmap of the levels of multiple immune-response-related proteins

**Supplementary Table 1. Maternal and newborn characteristics comparing between all twins and discordant twins**

| Characteristics | All twins (n=149) | Discordant twins (n=68) | $P^a$ |
|---|---|---|---|
| **Mothers** | | | |
| Age at delivery (years), mean±SD | 32.04±4.07 | 31.70±4.26 | 0.575[b] |
| <25, n (%) | 4 (2.68) | 2 (2.94) | 0.491 |
| 25-29, n (%) | 45 (30.20) | 26 (38.24) | |
| 30-34, n (%) | 69 (46.31) | 24 (35.29) | |
| ≥35, n (%) | 31 (20.81) | 16 (23.53) | |
| Educational level, n (%) | | | |
| Junior high school and below | 21 (14.09) | 10 (14.71) | 0.876 |
| Senior high/Vocational school | 50 (33.56) | 18 (26.47) | |
| University and above | 78 (52.35) | 40 (58.82) | |
| Pre-pregnancy BMI, mean±SD | 21.66±3.14 | 21.55±2.86 | 0.790[b] |
| <18.5, n (%) | 19 (12.75) | 6 (8.82) | 0.706 |
| 18.5-23.9, n (%) | 98 (65.77) | 50 (73.53) | |
| 24-27.9, n (%) | 26 (17.45) | 10 (14.71) | |
| ≥28, n (%) | 6 (4.03) | 2 (2.94) | |
| Mode of conception, n (%) | | | |
| Naturally conceived | 70 (46.98) | 26 (38.24) | 0.291 |
| IVF-ET | 79 (53.02) | 42 (61.76) | |
| Gravidity, n (%) | | | |
| 1 | 60 (40.27) | 26 (38.24) | 0.747 |
| 2 | 45 (30.20) | 24 (35.29) | |
| ≥3 | 44 (29.53) | 18 (26.47) | |
| Vaginal bleeding during early pregnancy, n (%) | | | |
| No | 120 (80.54) | 54 (79.41) | 0.993 |
| Yes | 29 (19.46) | 14 (20.59) | |
| Diabetes, n (%) | | | |
| No | 89 (59.73) | 44 (64.71) | 0.584 |
| Pregestational diabetes | 0 (0.00) | 0 (0.00) | |
| Gestational diabetes | 60 (40.27) | 24 (35.29) | |
| Hypertension, n (%) | | | |
| No | 106 (71.14) | 52 (76.47) | 0.513 |
| Pregestational hypertension | 0 (0.00) | 0 (0.00) | |
| Gestational hypertension | 43 (28.86) | 16 (23.53) | |
| Autoimmune diseases, n (%) | | | |
| No | 143 (95.97) | 64 (94.12) | 0.798 |
| Yes | 6 (4.03) | 4 (5.88) | |
| Gestational age (weeks), mean±SD | 36.37±1.16 | 36.54±1.07 | 0.292[b] |
| $<34^{+0}$, n (%) | 5 (3.36) | 2 (2.94) | 0.207 |
| $34^{+0}$-$36^{+6}$, n (%) | 80 (53.69) | 28 (41.18) | |
| $37^{+0}$-$38^{+6}$, n (%) | 64 (42.95) | 38 (55.88) | |
| $\geq 39^{+0}$, n (%) | 0 (0.00) | 0 (0.00) | |
| **Neonatal characteristics** | | | |
| Sex, n (%) | | | |
| Male | 76 (51.01) | 35 (51.47) | >0.999 |
| Female | 73 (48.99) | 33 (48.53) | |
| Birth weight (grams), mean±SD | 2,404.50±411.44 | 2,466.03±346.75 | 0.292[b] |
| <2000, n (%) | 28 (18.79) | 8 (11.76) | 0.567 |
| 2000-2499, n (%) | 98 (65.77) | 50 (73.53) | |
| 2500-2999, n (%) | 67 (44.97) | 32 (47.06) | |
| ≥3000, n (%) | 9 (6.04) | 6 (8.82) | |

SD, standard deviation; IVF-ET: In vitro fertilization and embryo transfer.

[a] Pearson's Chi-squared test or Fisher's exact test unless stated otherwise.

[b] Independent sample t-test.

**Supplementary Table 2. Normalized protein expression values of immune-response-related proteins between neonates without and with infection**

| Proteins | All twins (n=149) | | | Discordant twins (n=68) | | |
|---|---|---|---|---|---|---|
| | Without neonatal infection (n=89) | With neonatal infection (n=60) | *P* [1] | Without neonatal infection(n=34) | With neonatal infection (n=34) | *P* [2] |
| PPP1R9B | 1.58 (1.24-1.97) | 1.45 (1.12-1.92) | 0.199 | 1.53 (1.26-2.03) | 1.54 (1.17-1.94) | 0.748 |
| GLB1 | 0.00 (-0.21-0.40) | 0.04 (-0.21-0.30) | 0.485 | 0.05 (-0.20-0.39) | 0.01 (-0.40-0.26) | 0.301 |
| PSIP1 | 2.63 (2.23-3.38) | 2.55 (2.15-3.05) | 0.527 | 2.53 (2.25-3.11) | 2.66 (2.46-3.33) | 0.326 |
| ZBTB16 | 0.67 (0.30-1.09) | 0.43 (0.14-0.86) | 0.125 | 0.75 (0.49-1.08) | 0.40 (0.17-0.71) | 0.087 |
| IRAK4 | 2.31 (1.91-2.78) | 2.17 (1.72-2.74) | 0.266 | 2.46 (1.81-2.81) | 2.02 (1.67-2.63) | 0.081 |
| TPSAB1 | 4.67 (4.07-4.90) | 4.66 (4.07-4.98) | 0.946 | 4.28 (3.89-4.76) | 4.57 (4.09-4.81) | 0.510 |
| HCLS1 | 3.96 (3.58-4.50) | 3.77 (3.45-4.17) | 0.077 | 4.04 (3.65-4.36) | 3.82 (3.56-4.49) | 0.748 |
| CNTNAP2 | 2.58 (2.16-2.89) | 2.63 (2.41-2.99) | 0.058 | 2.67 (2.34-3.08) | 2.58 (2.38-2.83) | 0.761 |
| CLEC4G | 2.01 (1.81-2.23) | 2.02 (1.77-2.21) | 0.848 | 2.08 (1.84-2.24) | 1.98 (1.75-2.16) | 0.543 |
| IRF9 | 0.50 (0.26-0.96) | 0.58 (0.23-0.95) | 0.998 | 0.52 (0.39-0.97) | 0.57 (0.31-0.94) | 0.636 |
| EDAR | **1.87 (1.55-2.20)** | **1.61 (1.42-1.98)** | **0.023** | 1.73 (1.52-1.97) | 1.61 (1.48-2.09) | 0.919 |
| IL6 | 1.23 (0.86-1.56) | 1.04 (0.67-1.56) | 0.179 | 1.17 (0.76-1.39) | 1.14 (0.66-1.43) | 0.698 |
| DGKZ | 0.17 (-0.25-0.46) | 0.00 (-0.38-0.26) | 0.096 | 0.04 (-0.34-0.47) | -0.04 (-0.53-0.17) | 0.407 |
| CLEC4C | 2.92 (2.62-3.18) | 2.88 (2.59-3.19) | 0.827 | 2.75 (2.52-3.09) | 2.88 (2.58-3.19) | 0.648 |
| IRAK1 | 0.99 (0.65-1.32) | 0.91 (0.63-1.32) | 0.767 | 1.02 (0.68-1.26) | 0.94 (0.62-1.34) | 0.532 |
| CLEC4A | 2.31 (2.08-2.55) | 2.37 (2.14-2.57) | 0.385 | 2.31 (2.03-2.50) | 2.29 (2.15-2.58) | 0.235 |
| PRDX1 | 4.43 (3.76-5.40) | 4.23 (3.63-5.04) | 0.393 | 4.48 (3.91-5.19) | 4.30 (3.83-5.00) | 0.800 |
| PRDX3 | -0.13 (-0.41-0.24) | -0.10 (-0.35-0.18) | 0.958 | 0.02 (-0.28-0.41) | -0.17 (-0.44-0.06) | 0.004 |
| FGF2 | 0.86 (0.38-1.48) | 0.63 (0.37-1.04) | 0.193 | 0.71 (0.38-1.37) | 0.63 (0.41-1.26) | 0.577 |
| PRDX5 | 3.90 (3.29-4.74) | 3.85 (3.37-4.58) | 0.931 | 4.30 (3.71-4.78) | 4.03 (3.48-4.62) | 0.787 |
| DPP10 | 0.89 (0.63-1.25) | 1.00 (0.67-1.26) | 0.424 | 0.94 (0.71-1.39) | 0.78 (0.59-1.27) | 0.235 |
| TRIM5 | 2.06 (1.64-2.46) | 1.90 (1.64-2.36) | 0.325 | 2.08 (1.66-2.36) | 1.83 (1.61-2.32) | 0.163 |
| DCTN1 | 3.56 (3.18-4.38) | 3.47 (2.77-4.09) | 0.093 | 3.55 (3.30-4.29) | 3.74 (2.99-4.18) | 0.787 |
| ITGA6 | 2.13 (1.89-2.55) | 2.14 (1.93-2.38) | 0.504 | 2.10 (1.92-2.51) | 2.08 (1.92-2.37) | 0.467 |
| CDSN | 5.22 (4.96-5.47) | 5.18 (4.81-5.50) | 0.776 | 5.17 (4.98-5.37) | 5.12 (4.67-5.38) | 0.360 |
| GALNT3 | 4.58 (4.20-4.81) | 4.40 (4.19-4.76) | 0.362 | **4.65 (4.32-4.91)** | **4.39 (4.22-4.62)** | **0.031** |
| FXYD5 | 1.83 (1.61-2.11) | 1.85 (1.58-2.07) | 0.689 | 1.89 (1.66-2.08) | 1.81 (1.54-2.00) | 0.577 |
| TRAF2 | 2.98 (2.78-3.57) | 3.04 (2.71-3.43) | 0.545 | 2.96 (2.82-3.32) | 3.05 (2.76-3.47) | 0.774 |
| TRIM21 | 3.29 (2.97-3.71) | 3.22 (2.93-3.55) | 0.603 | 3.29 (3.13-3.53) | 3.28 (3.14-3.71) | 0.879 |
| LILRB4 | 2.91 (2.71-3.13) | 2.95 (2.69-3.08) | 0.857 | 2.85 (2.66-3.04) | 2.89 (2.70-3.02) | 0.648 |
| NTF4 | 2.99 (2.75-3.20) | 3.06 (2.85-3.27) | 0.358 | 2.97 (2.76-3.20) | 3.02 (2.79-3.28) | 0.698 |
| KRT19 | 3.99 (3.69-4.32) | 3.95 (3.63-4.34) | 0.931 | 3.88 (3.69-4.18) | 3.96 (3.70-4.30) | 0.278 |
| ITM2A | 2.52 (2.33-2.73) | 2.61 (2.41-2.85) | 0.085 | 2.58 (2.42-2.84) | 2.55 (2.37-2.94) | 0.761 |
| HNMT | 8.36 (7.91-8.95) | 8.31 (7.72-8.84) | 0.466 | 8.41 (7.83-8.93) | 8.43 (7.71-8.86) | 0.467 |
| CCL11 | 5.36 (4.92-5.68) | 5.14 (4.86-5.63) | 0.310 | 5.36 (5.15-5.71) | 5.15 (4.89-5.64) | 0.221 |
| MILR1 | 2.60 (2.34-2.95) | 2.58 (2.22-2.92) | 0.700 | 2.55 (2.31-2.98) | 2.51 (2.14-2.86) | 0.153 |
| EGLN1 | 1.49 (1.11-1.94) | 1.48 (1.03-2.00) | 0.424 | 1.51 (1.12-1.88) | 1.54 (1.04-1.94) | 0.736 |
| NFATC3 | -0.77 (-0.88--0.52) | -0.78 (-0.99--0.50) | 0.495 | -0.77 (-0.86--0.53) | -0.79 (-0.99--0.50) | 1.000 |
| LY75 | 0.62 (0.47-0.84) | 0.72 (0.43-0.91) | 0.229 | 0.80 (0.57-0.91) | 0.65 (0.35-0.87) | 0.078 |
| EIF5A | -0.09 (-0.43-0.24) | -0.04 (-0.32-0.29) | 0.692 | 0.11 (-0.36-0.32) | -0.04 (-0.30-0.27) | 0.879 |
| EIF4G1 | 4.38 (3.66-4.88) | 4.16 (3.60-4.77) | 0.350 | 4.38 (3.71-4.62) | 4.25 (3.68-4.74) | 0.826 |
| CD28 | 1.74 (1.46-1.95) | 1.74 (1.60-1.98) | 0.372 | 1.74 (1.51-1.99) | 1.71 (1.58-1.90) | 0.602 |
| PTH1R | 5.15 (5.03-5.34) | 5.19 (4.95-5.33) | 0.597 | **5.23 (5.05-5.34)** | **5.12 (4.87-5.26)** | **0.043** |
| BIRC2 | 0.06 (-0.18-0.28) | 0.07 (-0.16-0.23) | 0.830 | -0.02 (-0.28-0.27) | -0.02 (-0.17-0.18) | 0.589 |
| HSD11B1 | -0.87 (-1.17--0.59) | -0.86 (-1.16--0.55) | 0.872 | -0.79 (-1.08--0.53) | -0.88 (-1.15--0.57) | 0.317 |
| NF2 | 0.41 (0.11-0.89) | 0.29 (0.08-0.85) | 0.374 | 0.42 (0.22-0.78) | 0.34 (0.08-0.88) | 0.906 |
| PLXNA4 | 3.35 (3.00-4.37) | 3.20 (2.89-3.58) | 0.182 | 3.30 (3.00-4.18) | 3.29 (2.90-4.16) | 0.774 |
| SH2B3 | 1.60 (1.17-2.00) | 1.54 (1.06-1.95) | 0.334 | 1.63 (1.20-1.94) | 1.48 (1.08-1.75) | 0.478 |
| FCRL3 | -0.13 (-0.29-0.05) | -0.16 (-0.29-0.09) | 0.761 | -0.04 (-0.19-0.09) | -0.16 (-0.30-0.03) | 0.256 |
| CKAP4 | 7.82 (7.59-8.04) | 7.84 (7.56-7.96) | 0.619 | 7.79 (7.59-8.03) | 7.80 (7.46-7.92) | 0.343 |
| JUN | 1.85 (1.52-2.17) | 1.81 (1.56-2.13) | 0.940 | 1.94 (1.60-2.28) | 1.83 (1.59-2.12) | 0.168 |
| HEXIM1 | 3.91 (3.52-4.43) | 3.83 (3.56-4.22) | 0.466 | 3.90 (3.62-4.41) | 3.92 (3.58-4.21) | 0.800 |
| CLEC4D | 2.84 (2.41-3.31) | 2.91 (2.60-3.41) | 0.442 | 3.00 (2.63-3.34) | 2.97 (2.75-3.47) | 0.987 |
| PRKCQ | 0.97 (0.70-1.43) | 1.02 (0.66-1.30) | 0.909 | 0.89 (0.66-1.25) | 1.02 (0.67-1.26) | 0.636 |
| MGMT | 4.24 (3.64-4.97) | 4.05 (3.62-4.61) | 0.358 | 4.32 (3.84-4.74) | 4.32 (3.75-4.61) | 0.933 |
| TREM1 | 2.12 (1.81-2.45) | 2.09 (1.94-2.37) | 0.712 | 2.08 (1.88-2.34) | 2.20 (1.98-2.37) | 0.235 |
| CXADR | 3.97 (3.77-4.22) | 4.01 (3.80-4.27) | 0.424 | 4.11 (3.80-4.32) | 3.98 (3.74-4.26) | 0.407 |
| IL10 | 3.27 (2.97-3.57) | 3.19 (2.91-3.61) | 0.449 | 3.16 (2.98-3.50) | 3.03 (2.73-3.43) | 0.109 |
| SRPK2 | 0.92 (0.58-1.24) | 0.83 (0.59-1.17) | 0.269 | 0.91 (0.72-1.16) | 0.83 (0.64-1.22) | 0.946 |
| KLRD1 | 4.44 (4.08-4.74) | 4.40 (4.09-4.69) | 0.720 | 4.40 (3.98-4.59) | 4.43 (3.94-4.68) | 0.309 |
| BACH1 | 1.79 (1.59-2.12) | 1.83 (1.60-2.07) | 0.900 | 1.77 (1.61-2.03) | 1.82 (1.60-2.14) | 0.879 |
| PIK3AP1 | 2.97 (2.58-3.43) | 2.84 (2.53-3.35) | 0.519 | 2.98 (2.67-3.36) | 2.99 (2.63-3.38) | 0.987 |
| SPRY2 | 2.98 (2.52-3.37) | 2.87 (2.23-3.18) | 0.104 | 2.77 (2.35-3.21) | 2.85 (2.17-3.17) | 0.813 |

| | | | | | | |
|---|---|---|---|---|---|---|
| STC1 | 5.61 (5.50-5.73) | 5.59 (5.42-5.70) | 0.499 | 5.60 (5.45-5.72) | 5.58 (5.44-5.70) | 0.787 |
| ARNT | 1.84 (1.43-2.13) | 1.81 (1.47-2.21) | 0.800 | 1.88 (1.54-2.22) | 1.63 (1.46-2.01) | 0.388 |
| FAM3B | 5.88 (5.64-6.05) | 5.88 (5.67-6.11) | 0.630 | 5.74 (5.56-5.99) | 5.73 (5.62-5.90) | 0.774 |
| SH2D1A | **2.16 (1.83-2.61)** | **1.97 (1.66-2.27)** | **0.018** | 2.08 (1.87-2.34) | 1.97 (1.68-2.27) | 0.748 |
| ICA1 | 1.76 (1.28-2.09) | 1.52 (1.20-2.00) | 0.108 | 1.73 (1.25-2.06) | 1.42 (1.10-1.65) | 0.050 |
| DFFA | 5.25 (4.82-5.72) | 5.11 (4.68-5.59) | 0.184 | 5.21 (4.87-5.57) | 5.11 (4.73-5.47) | 0.813 |
| DCBLD2 | 7.31 (7.13-7.43) | 7.28 (7.17-7.42) | 0.818 | 7.33 (7.13-7.43) | 7.21 (7.16-7.39) | 0.360 |
| FCRL6 | **1.86 (1.62-2.12)** | **1.99 (1.80-2.12)** | **0.039** | 1.97 (1.76-2.14) | 1.97 (1.75-2.15) | 0.840 |
| NCR1 | 3.12 (2.85-3.37) | 2.99 (2.83-3.36) | 0.402 | 3.08 (2.84-3.36) | 2.93 (2.77-3.24) | 0.084 |
| CXCL12 | 0.20 (-0.11-0.53) | 0.24 (-0.10-0.59) | 0.885 | 0.28 (0.05-0.79) | 0.14 (-0.19-0.42) | 0.087 |
| AREG | 4.16 (3.75-4.55) | 4.18 (3.62-4.85) | 0.791 | 4.07 (3.69-4.52) | 4.13 (3.63-4.61) | 0.879 |
| IFNLR1 | 1.99 (1.84-2.28) | 1.92 (1.68-2.17) | 0.141 | 1.94 (1.81-2.12) | 1.82 (1.58-2.09) | 0.121 |
| DAPP1 | 2.17 (1.79-2.60) | 2.07 (1.73-2.47) | 0.290 | 2.20 (1.90-2.59) | 2.18 (1.91-2.73) | 0.685 |
| PADI2 | -0.24 (-0.43--0.03) | -0.36 (-0.50--0.00) | 0.293 | -0.16 (-0.39-0.12) | -0.41 (-0.50--0.23) | 0.033 |
| SIT1 | 3.91 (3.32-4.81) | 4.02 (3.12-4.56) | 0.396 | 3.82 (3.30-4.48) | 4.08 (3.26-4.53) | 0.532 |
| MASP1 | 0.46 (0.24-0.60) | 0.51 (0.36-0.69) | 0.100 | 0.45 (0.19-0.58) | 0.49 (0.35-0.59) | 0.326 |
| LAMP3 | 3.61 (3.41-3.82) | 3.57 (3.40-3.85) | 0.885 | 3.55 (3.36-3.81) | 3.53 (3.36-3.81) | 0.543 |
| CLEC7A | 2.30 (2.06-2.53) | 2.35 (2.11-2.59) | 0.398 | 2.39 (2.20-2.63) | 2.35 (2.08-2.61) | 0.309 |
| CLEC6A | 2.10 (1.77-2.42) | 2.04 (1.73-2.42) | 0.821 | 2.14 (1.97-2.41) | 1.91 (1.57-2.36) | 0.072 |
| DDX58 | **2.41 (1.99-2.69)** | **2.03 (1.82-2.40)** | **0.010** | 2.28 (1.96-2.62) | 2.17 (1.85-2.45) | 0.326 |
| IL12RB1 | 1.27 (1.03-1.51) | 1.17 (1.04-1.44) | 0.581 | 1.24 (0.89-1.51) | 1.13 (1.02-1.34) | 0.612 |
| TANK | 2.70 (2.32-3.04) | 2.66 (2.25-2.98) | 0.470 | 2.67 (2.33-3.00) | 2.66 (2.26-2.91) | 0.532 |
| ITGA11 | **0.97 (0.53-1.22)** | **1.13 (0.83-1.44)** | **0.033** | 0.95 (0.41-1.22) | 1.09 (0.82-1.37) | 0.326 |
| KPNA1 | -0.87 (-1.24--0.38) | -0.66 (-1.21--0.37) | 0.310 | -0.84 (-1.18--0.30) | -0.66 (-1.24--0.33) | 0.919 |
| LAG3 | 0.66 (0.44-0.80) | 0.57 (0.30-0.83) | 0.211 | **0.67 (0.47-0.81)** | **0.45 (0.27-0.66)** | **0.031** |
| IL5 | 0.40 (0.04-0.81) | 0.31 (-0.01-0.74) | 0.269 | 0.54 (-0.08-1.08) | 0.21 (-0.04-0.52) | 0.109 |
| CD83 | 2.15 (1.93-2.31) | 2.13 (1.93-2.38) | 0.700 | 2.17 (1.92-2.25) | 2.07 (1.92-2.27) | 0.933 |
| ITGB6 | 3.44 (3.21-3.66) | 3.41 (3.25-3.70) | 0.468 | 3.51 (3.26-3.61) | 3.37 (3.16-3.69) | 0.397 |
| BTN3A2 | 2.86 (2.67-3.07) | 2.84 (2.62-3.15) | 0.918 | 2.87 (2.75-3.11) | 2.78 (2.61-3.12) | 0.388 |

Median (Interquartile range)

[1]Wilcoxon rank-sum test.

[2]Wilcoxon signed-rank test.

**Supplementary Table 3. Association between immune-response-related proteins levels and neonatal infection in twins**

| Proteins | All twins (n=149) | | Discordant twins (n=68) | |
|---|---|---|---|---|
| | *OR* (95%*CI*) | *OR* (95%*CI*)[a] | *OR* (95%*CI*) | *OR* (95%*CI*)[b] |
| PPP1R9B | 0.68 (0.38-1.21) | 0.64 (0.32-1.29) | 0.83 (0.36-1.94) | 1.21 (0.41-3.61) |
| GLB1 | 0.73 (0.39-1.39) | 0.66 (0.30-1.42) | 0.63 (0.26-1.53) | 0.65 (0.26-1.64) |
| PSIP1 | 0.84 (0.55-1.27) | 0.80 (0.46-1.39) | 1.60 (0.72-3.55) | **3.49 (1.07-11.44)** |
| ZBTB16 | 0.71 (0.42-1.20) | **0.47 (0.23-0.95)** | 0.61 (0.26-1.46) | 1.02 (0.31-3.41) |
| IRAK4 | 0.79 (0.52-1.21) | 0.74 (0.44-1.25) | 0.85 (0.48-1.51) | 1.08 (0.55-2.12) |
| TPSAB1 | 0.93 (0.60-1.44) | 1.03 (0.62-1.71) | 1.64 (0.70-3.84) | 1.74 (0.66-4.56) |
| HCLS1 | 0.75 (0.50-1.13) | 0.70 (0.43-1.15) | 1.16 (0.54-2.48) | 1.71 (0.68-4.30) |
| CNTNAP2 | **2.10 (1.06-4.13)** | **2.34 (1.04-5.28)** | 0.68 (0.14-3.17) | 0.84 (0.13-5.45) |
| CLEC4G | 1.12 (0.52-2.39) | 1.81 (0.65-5.03) | 0.38 (0.08-1.92) | 0.32 (0.05-2.08) |
| IRF9 | 0.93 (0.57-1.54) | 0.95 (0.50-1.80) | 1.12 (0.43-2.89) | 2.09 (0.55-8.00) |
| EDAR | 0.60 (0.32-1.12) | 0.57 (0.27-1.19) | 1.12 (0.52-2.41) | 1.60 (0.63-4.06) |
| IL6 | 0.75 (0.44-1.28) | 0.68 (0.35-1.32) | 1.19 (0.46-3.05) | 0.90 (0.25-3.18) |
| DGKZ | 0.63 (0.31-1.27) | 0.57 (0.24-1.36) | 0.75 (0.30-1.91) | 0.95 (0.25-3.56) |
| CLEC4C | 1.00 (0.48-2.07) | 1.24 (0.51-3.02) | 1.61 (0.53-4.92) | 1.36 (0.33-5.58) |
| IRAK1 | 0.94 (0.54-1.64) | 0.96 (0.50-1.86) | 1.10 (0.51-2.38) | 1.45 (0.60-3.55) |
| CLEC4A | 1.55 (0.57-4.17) | 2.25 (0.68-7.47) | 3.29 (0.57-19.11) | 4.12 (0.47-36.34) |
| PRDX1 | 0.90 (0.70-1.17) | 0.92 (0.68-1.24) | 1.14 (0.73-1.78) | 1.46 (0.84-2.52) |
| PRDX3 | 0.96 (0.55-1.65) | 0.76 (0.37-1.54) | **0.22 (0.05-0.95)** | 0.21 (0.03-1.44) |
| FGF2 | 0.81 (0.56-1.17) | 0.85 (0.54-1.32) | 1.07 (0.60-1.92) | 2.25 (0.91-5.58) |
| PRDX5 | 0.95 (0.72-1.25) | 0.93 (0.66-1.32) | 1.04 (0.61-1.78) | 1.28 (0.66-2.49) |
| DPP10 | 1.27 (0.73-2.23) | 1.75 (0.89-3.45) | 0.31 (0.05-1.78) | 0.40 (0.03-4.83) |
| TRIM5 | 0.77 (0.46-1.30) | 0.66 (0.35-1.25) | 0.86 (0.39-1.89) | 1.12 (0.42-2.95) |
| DCTN1 | 0.78 (0.56-1.10) | 0.77 (0.51-1.16) | 1.03 (0.56-1.88) | 1.51 (0.71-3.19) |
| ITGA6 | 0.72 (0.38-1.36) | 0.86 (0.40-1.89) | 0.67 (0.24-1.89) | 1.16 (0.34-3.99) |
| CDSN | 0.80 (0.43-1.49) | 0.64 (0.31-1.33) | 0.34 (0.08-1.50) | 0.22 (0.03-1.47) |
| GALNT3 | 0.82 (0.45-1.52) | 1.04 (0.50-2.16) | 0.41 (0.13-1.29) | 0.46 (0.10-2.10) |
| FXYD5 | 1.20 (0.62-2.31) | 1.01 (0.46-2.22) | 0.93 (0.31-2.74) | 1.66 (0.41-6.62) |
| TRAF2 | 0.89 (0.58-1.36) | 0.86 (0.51-1.44) | 1.46 (0.65-3.27) | 2.51 (0.89-7.09) |
| TRIM21 | 0.97 (0.60-1.54) | 1.22 (0.69-2.17) | 1.38 (0.61-3.13) | 1.93 (0.74-4.99) |
| LILRB4 | 1.50 (0.61-3.69) | 1.42 (0.47-4.28) | 2.83 (0.33-23.94) | 2.12 (0.15-30.78) |
| NTF4 | 1.47 (0.52-4.14) | 1.71 (0.47-6.22) | 1.01 (0.23-4.36) | 0.66 (0.11-3.74) |
| KRT19 | 0.90 (0.53-1.51) | 0.80 (0.45-1.44) | 2.21 (0.64-7.69) | 2.91 (0.59-14.25) |
| ITM2A | 2.32 (0.99-5.47) | 2.00 (0.77-5.17) | 1.57 (0.41-6.01) | 0.96 (0.20-4.53) |
| HNMT | 0.83 (0.57-1.20) | 0.78 (0.49-1.22) | 0.89 (0.44-1.83) | 0.97 (0.37-2.53) |
| CCL11 | 0.84 (0.48-1.46) | 1.21 (0.61-2.42) | 0.65 (0.18-2.32) | 1.01 (0.23-4.42) |
| MILR1 | 0.86 (0.45-1.64) | 0.73 (0.32-1.67) | 0.42 (0.12-1.51) | 0.29 (0.06-1.54) |
| EGLN1 | 0.79 (0.51-1.21) | 0.78 (0.46-1.31) | 1.08 (0.48-2.41) | 1.65 (0.63-4.36) |
| NFATC3 | 0.75 (0.33-1.74) | 0.60 (0.22-1.65) | 1.46 (0.26-8.25) | 2.86 (0.36-23.06) |
| LY75 | 2.09 (0.72-6.04) | 3.09 (0.84-11.36) | 0.17 (0.02-1.28) | 0.08 (0.01-1.25) |
| EIF5A | 1.24 (0.61-2.52) | 2.09 (0.82-5.30) | 0.82 (0.26-2.53) | 0.77 (0.21-2.86) |
| EIF4G1 | 0.89 (0.63-1.25) | 0.84 (0.55-1.27) | 1.14 (0.60-2.15) | 1.47 (0.69-3.11) |
| CD28 | 1.52 (0.57-4.02) | 1.33 (0.42-4.21) | 0.67 (0.16-2.86) | 1.01 (0.18-5.66) |
| PTH1R | 0.50 (0.15-1.69) | 0.48 (0.11-2.05) | 0.14 (0.02-1.08) | 0.12 (0.01-1.36) |
| BIRC2 | 0.96 (0.41-2.26) | 1.05 (0.38-2.95) | 0.78 (0.23-2.69) | 1.19 (0.26-5.53) |
| HSD11B1 | 1.06 (0.48-2.34) | 1.41 (0.53-3.74) | 0.50 (0.13-1.93) | 0.65 (0.11-3.79) |
| NF2 | 0.79 (0.47-1.31) | 0.84 (0.47-1.51) | 0.96 (0.48-1.93) | 1.61 (0.63-4.09) |
| PLXNA4 | 0.84 (0.60-1.18) | 0.72 (0.48-1.09) | 1.05 (0.66-1.66) | 1.20 (0.69-2.09) |
| SH2B3 | 0.80 (0.51-1.25) | 0.77 (0.45-1.33) | 0.82 (0.44-1.53) | 1.08 (0.47-2.49) |
| FCRL3 | 1.17 (0.41-3.32) | 1.00 (0.29-3.42) | 0.40 (0.08-1.89) | 0.34 (0.05-2.32) |
| CKAP4 | 0.85 (0.34-2.13) | 0.56 (0.18-1.76) | 0.45 (0.10-2.09) | 0.45 (0.07-2.71) |
| JUN | 1.04 (0.57-1.90) | 1.20 (0.60-2.42) | 0.65 (0.25-1.70) | 0.95 (0.35-2.61) |
| HEXIM1 | 0.88 (0.59-1.34) | 0.90 (0.55-1.47) | 1.23 (0.58-2.61) | 1.73 (0.72-4.17) |
| CLEC4D | 1.26 (0.76-2.08) | 1.50 (0.81-2.78) | 1.09 (0.44-2.70) | 1.27 (0.44-3.66) |
| PRKCQ | 0.99 (0.60-1.62) | 0.99 (0.56-1.73) | 1.01 (0.53-1.94) | 1.17 (0.55-2.46) |
| MGMT | 0.85 (0.61-1.19) | 0.78 (0.52-1.18) | 1.21 (0.68-2.17) | 1.77 (0.79-3.99) |
| TREM1 | 1.18 (0.56-2.48) | 1.93 (0.77-4.82) | 3.41 (0.58-20.17) | 9.92 (0.87-113.44) |
| CXADR | 1.79 (0.72-4.44) | 1.99 (0.67-5.93) | 0.32 (0.04-2.54) | 0.21 (0.01-5.91) |
| IL10 | 0.96 (0.66-1.39) | 0.82 (0.53-1.28) | 1.00 (0.57-1.76) | 1.01 (0.55-1.84) |
| SRPK2 | 0.66 (0.37-1.18) | 0.58 (0.28-1.19) | 1.36 (0.42-4.42) | 5.01 (0.87-28.95) |
| KLRD1 | 1.00 (0.49-2.03) | 1.05 (0.44-2.49) | 2.11 (0.54-8.24) | 1.53 (0.30-7.79) |
| BACH1 | 0.82 (0.51-1.35) | 0.82 (0.47-1.44) | 1.53 (0.61-3.86) | 2.66 (0.75-9.50) |
| PIK3AP1 | 0.86 (0.57-1.30) | 0.83 (0.50-1.39) | 1.16 (0.57-2.33) | 1.63 (0.71-3.71) |
| SPRY2 | 0.72 (0.47-1.12) | **0.54 (0.31-0.94)** | 1.15 (0.55-2.38) | 0.97 (0.41-2.32) |
| STC1 | 0.60 (0.11-3.32) | 1.26 (0.14-11.13) | 0.28 (0.01-9.80) | 0.25 (0.00-13.12) |
| ARNT | 1.23 (0.71-2.14) | 1.17 (0.60-2.28) | 0.88 (0.45-1.73) | 1.06 (0.43-2.60) |

| | | | | |
|---|---|---|---|---|
| FAM3B | 1.54 (0.51-4.65) | 1.69 (0.41-6.92) | 1.51 (0.23-10.03) | 1.50 (0.14-16.30) |
| SH2D1A | **0.53 (0.31-0.91)** | **0.28 (0.12-0.63)** | 0.94 (0.32-2.73) | 2.02 (0.51-8.01) |
| ICA1 | 0.68 (0.37-1.24) | 0.63 (0.30-1.33) | 0.47 (0.19-1.15) | 0.59 (0.17-2.03) |
| DFFA | 0.79 (0.52-1.20) | 0.78 (0.48-1.28) | 1.16 (0.56-2.41) | 1.65 (0.69-3.94) |
| DCBLD2 | 1.13 (0.30-4.26) | 1.99 (0.37-10.55) | 0.26 (0.02-3.92) | 0.77 (0.03-22.32) |
| FCRL6 | **3.83 (1.25-11.74)** | **5.72 (1.45-22.55)** | 0.72 (0.07-7.50) | 0.94 (0.08-11.61) |
| NCR1 | 0.83 (0.33-2.13) | 0.67 (0.21-2.15) | 0.26 (0.04-1.60) | 0.08 (0.01-1.03) |
| CXCL12 | 0.97 (0.52-1.83) | 0.91 (0.42-2.00) | 0.50 (0.21-1.19) | 0.60 (0.17-2.11) |
| AREG | 1.07 (0.73-1.57) | 1.07 (0.66-1.74) | 1.09 (0.47-2.56) | 0.83 (0.25-2.72) |
| IFNLR1 | 0.49 (0.18-1.33) | 0.45 (0.14-1.48) | 0.35 (0.08-1.50) | 0.35 (0.05-2.31) |
| DAPP1 | 0.74 (0.49-1.13) | 0.74 (0.44-1.24) | 0.83 (0.42-1.67) | 0.97 (0.41-2.31) |
| PADI2 | 0.95 (0.48-1.86) | 1.39 (0.62-3.10) | 0.32 (0.08-1.28) | 0.27 (0.04-2.05) |
| SIT1 | 0.83 (0.62-1.13) | 0.77 (0.53-1.14) | 1.16 (0.54-2.50) | 1.69 (0.62-4.59) |
| MASP1 | 2.99 (0.91-9.89) | **4.71 (1.11-19.94)** | 3.26 (0.36-29.63) | 3.62 (0.34-38.51) |
| LAMP3 | 0.95 (0.34-2.66) | 1.38 (0.39-4.93) | 0.82 (0.12-5.75) | 0.48 (0.04-6.27) |
| CLEC7A | 1.26 (0.50-3.21) | 1.53 (0.51-4.61) | 0.34 (0.05-2.49) | 0.35 (0.03-4.33) |
| CLEC6A | 0.99 (0.55-1.80) | 1.09 (0.54-2.20) | 0.37 (0.12-1.11) | 0.21 (0.04-1.00) |
| DDX58 | 0.62 (0.35-1.08) | 0.61 (0.31-1.23) | 1.03 (0.45-2.34) | 1.32 (0.47-3.71) |
| IL12RB1 | 0.87 (0.34-2.21) | 0.84 (0.28-2.53) | 0.70 (0.17-2.88) | 0.94 (0.14-6.29) |
| TANK | 0.78 (0.45-1.35) | 0.76 (0.39-1.49) | 0.80 (0.35-1.85) | 1.70 (0.42-6.94) |
| ITGA11 | **1.85 (1.00-3.40)** | **3.09 (1.38-6.96)** | 1.40 (0.63-3.07) | **5.50 (1.20-25.23)** |
| KPNA1 | 1.20 (0.80-1.80) | 1.38 (0.81-2.36) | 1.26 (0.65-2.44) | 1.20 (0.57-2.55) |
| LAG3 | 0.51 (0.17-1.52) | 0.46 (0.13-1.62) | 0.23 (0.05-1.10) | 0.15 (0.02-1.08) |
| IL5 | 0.76 (0.42-1.36) | 0.77 (0.39-1.52) | 0.53 (0.22-1.24) | 0.41 (0.11-1.55) |
| CD83 | 1.48 (0.47-4.65) | 1.34 (0.31-5.86) | 1.01 (0.16-6.23) | 0.76 (0.07-8.39) |
| ITGB6 | 1.50 (0.61-3.67) | 1.44 (0.47-4.47) | 0.33 (0.04-2.37) | 0.43 (0.05-3.93) |
| BTN3A2 | 0.95 (0.38-2.39) | 0.80 (0.27-2.38) | 0.40 (0.09-1.72) | 0.25 (0.04-1.51) |

OR: Odds ratios; CI: confidence interval.

[a]Adjusted for maternal age at delivery, educational level, pre-pregnancy BMI, mode of conception, gravidity, vaginal bleeding, gestational age, complications during pregnancy, as well as infant's sex and birth weight.

[b]Adjusted for infant's sex and birth weight.

**Supplementary Figure 1. Box plot for proteins levels between infant with or without neonatal infection**

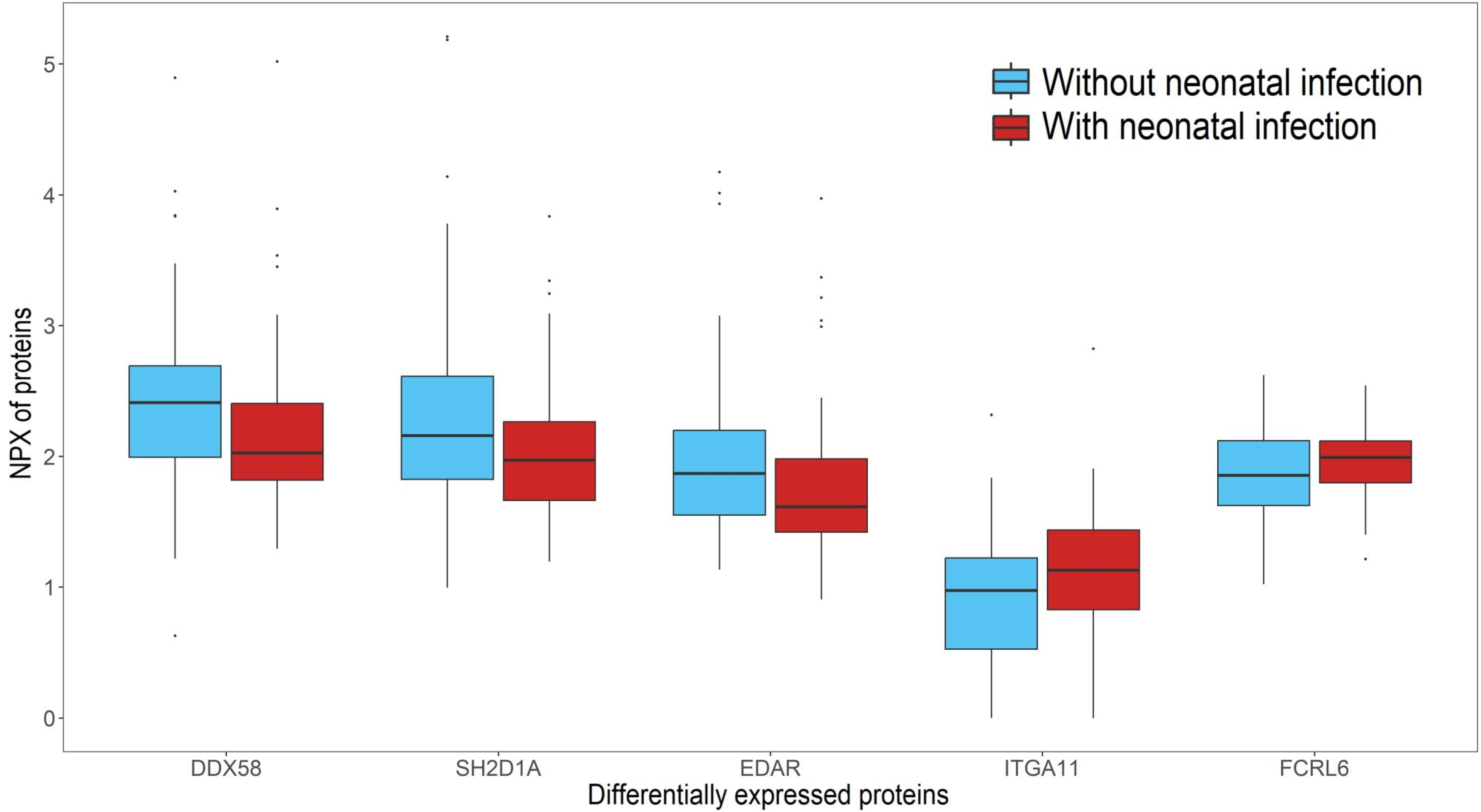

**Supplementary Figure 2. Cluster heatmap of the levels of multiple immune response-related proteins**

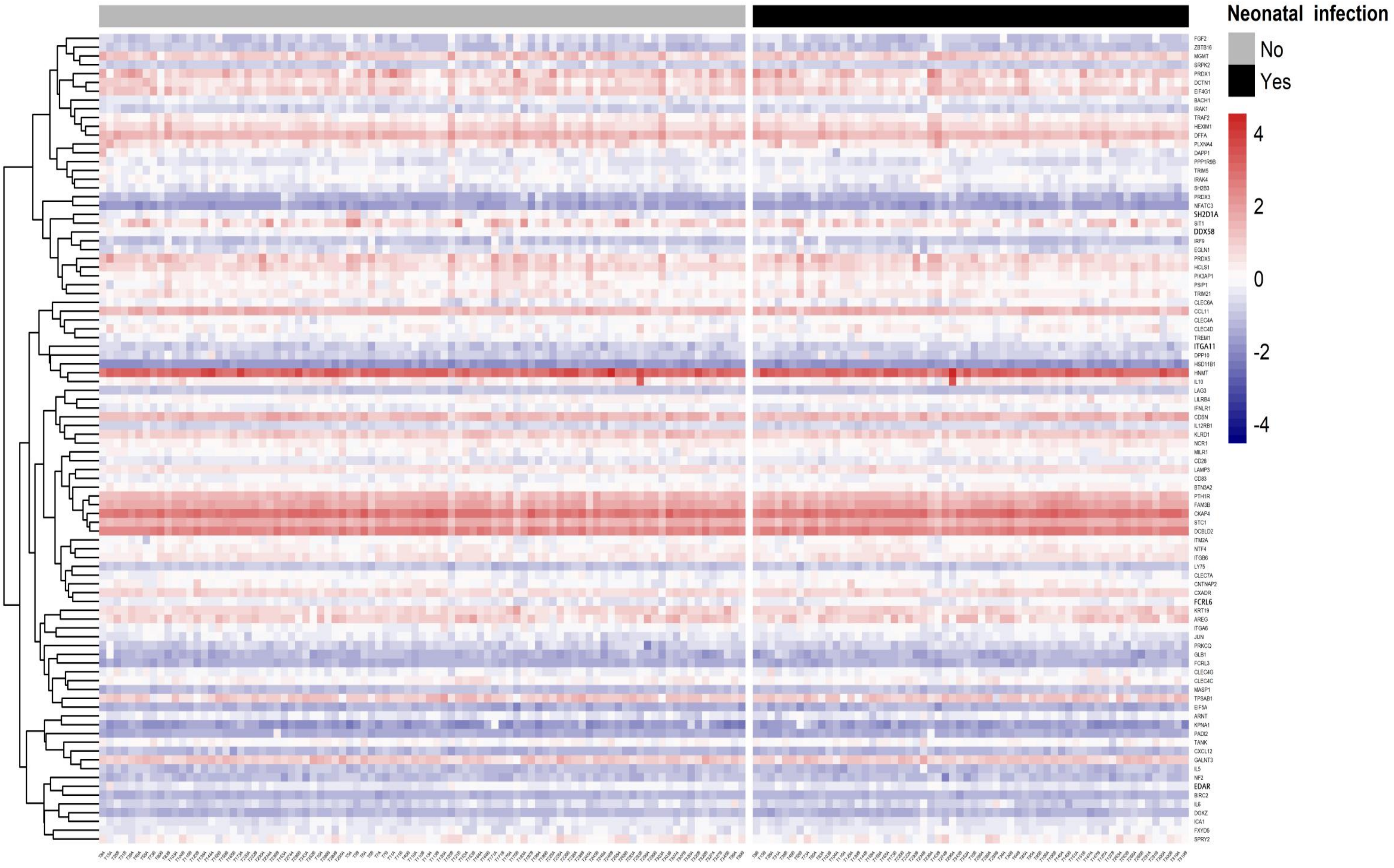

**Supplementary Figure 3. Correlation heatmap of the levels of multiple immune response-related proteins**

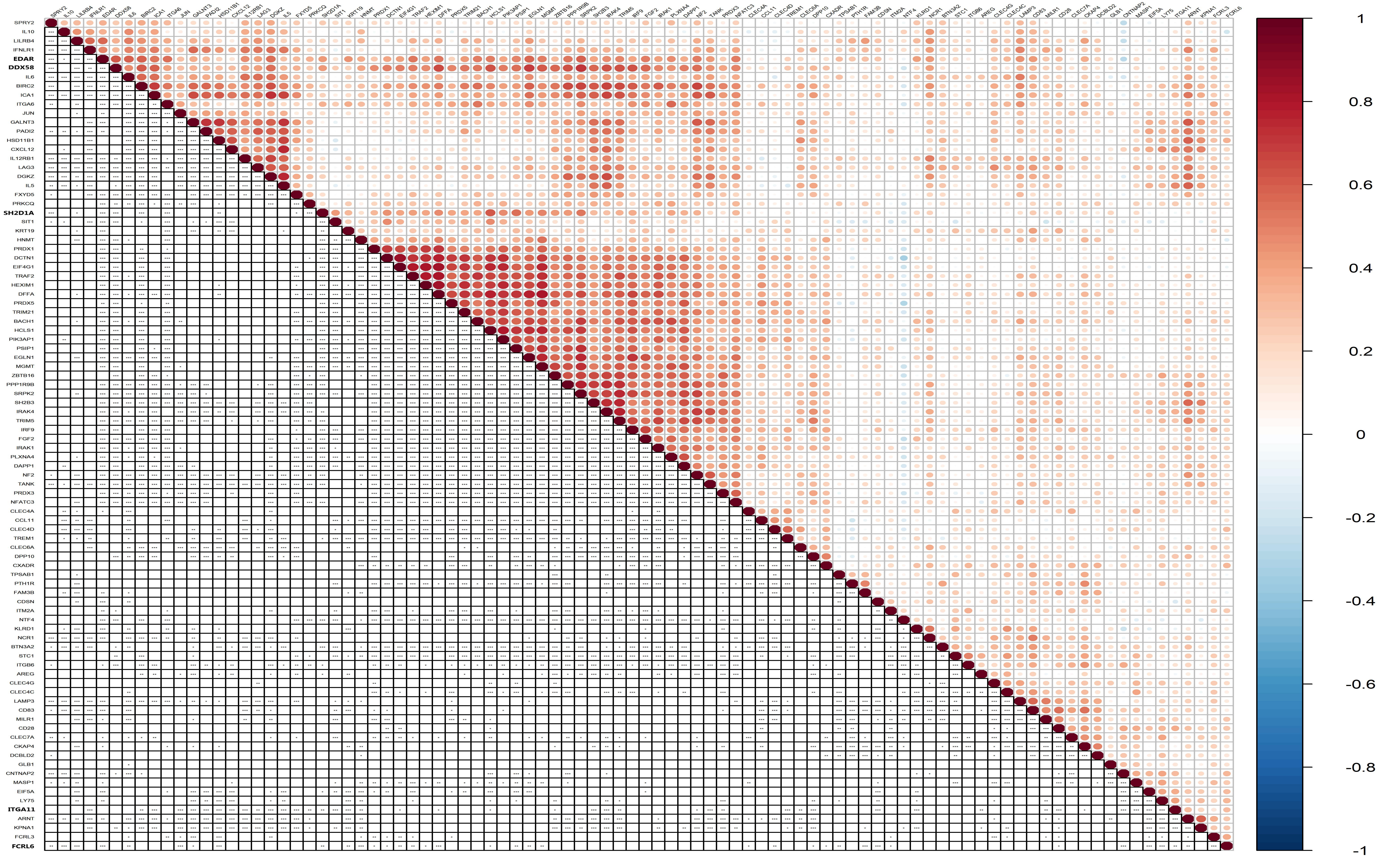
